# Examination of WHO/INRUD Core Drug Use Indicators at Public Primary Healthcare Centers in Kisii County, Kenya

**DOI:** 10.1101/2020.03.15.20036269

**Authors:** Faith A. Okalebo, Eric M. Guantai, Aggrey O. Nyabuti

## Abstract

**Background:** Irrational drug use is a global problem. However, the extent of the problem is higher in low-income countries. This study set out to assess and characterize drug use at the public primary healthcare centers (PPHCCs) in a rural county in Kenya, using the World Health Organization/ International Network for the Rational Use of Drugs (WHO/INRUD) core drug use indicators methodology.

**Methods:** Ten PPHCCs were randomly selected. From each PPHCC, ninety prescriptions from October to December 2018 were sampled and data extracted. Three-hundred (30 per PPHCC) patients and ten (1 per PPHCC) dispensers were also observed and interviewed. The WHO/INRUD core drug use indicators were used to assess the patterns of drug use.

**Results:** The average number of drugs per prescription was 2.9 (SD 0.5) (recommended: 1.6– 1.8), percentage of drugs prescribed by generic names was 27.7% (recommended: 100%); the percentage of prescriptions with an antibiotic was 84.8% (recommended: 20.0–26.8%), and with an injection prescribed was 24.9% (recommended: 13.4–24.1%). The percentage of prescribed drugs from the Kenya Essential Medicines List was 96.7% (recommended: 100%). The average consultation time was 4.1 min (SD 1.7) (recommended: ≥10 min), the average dispensing time was 131.5 sec (SD 41.5) (recommended: ≥90 sec), the percentage of drugs actually dispensed was 76.3% (recommended: 100%), the percentage of drugs adequately labeled was 22.6% (recommended: 100%) and percentage of patients with correct knowledge of dispensed drugs was 54.7% (recommended: 100%). Only 20% of the PPHCCs had a copy of KEML available, and 80% of the selected essential drugs assessed were available.

**Conclusion:** The survey shows irrational drug use practices, particularly polypharmacy, non-generic prescribing, overuse of antibiotics, short consultation time and inadequacy of drug labeling. Effective programs and activities promoting the rational use of drugs are the key interventions suggested at all the health facilities.

## Background

Drugs are very significant components of any healthcare system and should be used rationally. Rational drug use means that patients get medications suitable to their medical needs, in the right doses, for a suitable period of time, at the cheapest cost (1). Inappropriate use of drugs is an issue of concern with so many undesirable consequences such as the increased incidences of drug resistance, adverse drug reactions, cost of drug therapy, wastage of resources and reduced quality of drug therapy (2). Therefore irrational use of drugs leads to serious consequences, both in terms of healthcare and economics (3).

Irrational drug use may take many different forms, including poly-pharmacy, inappropriate use of injections and antibiotics, failure to comply with the standard treatment guidelines (STGs) while prescribing and inappropriate self-medication (4). Improvements in the manner in which drugs are used are very crucial in minimizing the morbidity and mortality associated with irrational drug use (5).

Drug use indicators have been established by the World Health Organization and the International Network for Rational Use of Drugs (WHO/INRUD) (6). They are broadly divided into two groups, namely core and complementary indicators. The core indicators have been pre-tested and standardized, and are grouped into three major categories namely prescribing, patient-care and facility-specific indicators (7). These drug use indicators are usually used in assessing drug use in out-patient facilities, where they provide measures of the optimal drug use as well as identify areas of deviations from the expected standards.

Primary healthcare (PHC) is a very crucial part of the healthcare system and is responsible for providing basic healthcare services. There is a six-level hierarchy of health facilities in the Kenyan health system, in ascending order they include: community services, dispensaries and clinics, health centers and nursing and maternity homes, sub-county hospitals, county referral hospitals and national referral hospitals and large private teaching hospitals. PHC services are mainly provided at the community services, dispensaries and clinics(10).

Irrational use of drugs can result in wastage of resources and widespread health hazards. A survey carried out at the health facilities of Southern Malawi showed that the country wasted its financial resources in the purchase of excessive drugs which ended up being used irrationally and quite a number expiring at the health facilities’ stores (11). In a study done in Jordan, the average number of drugs prescribed per encounter was higher against the WHO standards; there was a lower percentage of generic prescribing. The rest of the prescribing indicators including the injections prescribing, antibiotics prescribing, and prescribing from the essential medicines list were within the optimal range of values recommended by the WHO (12). Also, in a study carried out in Eritrea’s community pharmacies, it was found that the percentage of antibiotics being prescribed at the community pharmacies in Asmara was 53% which deviated significantly from the WHO recommended values. Furthermore, the percentage of encounters with injection was 7.8% lower than the WHO value. Patients’ age, gender and number of drugs were significantly associated with antibiotic prescribing (13).

Due to the complexity of drug use, it is important for it to be assessed so that problems may be identified and interventional strategies implemented so as to keep on the check the unsafe trends in drug utilization. Studies done in different parts of the world show that there are different drug use patterns, and a few such surveys have been carried out in Kenya.

Since no study of this kind has ever been conducted in Kisii County since the inception of devolution of healthcare in 2010, it was most likely that the County Government was wasting its resources on irrational drug use

This study, therefore, set out to use the WHO/INRUD core drug use indicators methodology to examine the patterns of drug use and the prevalence of irrational drug use at the public primary healthcare centers (level II and III facilities) in Kisii County, Kenya.

## METHODS

### Study area

The study was conducted at public primary health-care centers (PPHCCs) in Kisii County. This county in western Kenya has a total of 104 operational PPHCCs comprised of levels II (81) and III (23) facilities. The clientele of these centers is drawn from a population of about 1.2 million people from the entire county as well as the neighboring counties.

### Study design

The study was a hospital-based cross-sectional survey. Ten PPHCCs within the county were selected by a simple random sampling method. A survey was performed on patient prescriptions issued during the last quarter of 2018 (1^st^ October – 31^st^ December 2018). A total of 900 prescriptions (90 per PPHCC) were sampled by systematic random sampling. Patient-care and facility-specific surveys were conducted concurrently. For the patient-care survey, a total of 300 hundred patients (30 per PPHCC) who visited the facility during the survey period were recruited by convenient sampling as they waited to see the prescribing officer. Also, one dispenser from each PPHCC was recruited.

### Data collection

Prescription survey and patient-care survey data were collected by trained research assistants using standardized data collection forms. Data on patient-specific indicators were collected from participating patients by both direct observation and interviews as the patients moved from the prescribing area to the dispensing area. One dispenser from each of the selected PPHCC was interviewed to collect data on the key aspects of facility-specific indicators such as availability of copies of the Kenya Essential Medicines List (KEML) and availability of key drugs at the facility.

### Data analysis

Data collected in the specific indicator forms were entered into the – Epi Info™ version 7.2.2.16 (Centers for Disease Control and Prevention, US) and then exported to STATA version 14.2 (StataCorp, USA) for analysis. The data summarized using means, standard deviations, frequencies, and percentages. The ANOVA test was also used to test for differences between the PPHCCs. The core drug use indicators were also determined as described in guidelines for calculating the WHO/INRUD drug use indicators (14).

### Ethical considerations

Ethical approval to carry out the study was granted by the Kenyatta National Hospital - University of Nairobi Ethics and Research Committee (KNH/UoN-ERC) (Reference number: KNH – ERC/A/50). Permission to conduct the survey was also granted by the office of the Director for Health, Kisii County. Informed consent was obtained in writing from the patients, prescribers, and dispensers before conducting the interviews.

## RESULTS

The study was carried out at ten randomly sampled public primary healthcare centers (PPHCCs) in Kisii County - 5 levels II and 5 levels III health facilities. The total outpatient attendance of patients at the selected PPHCCs in the last quarter of 2018 39,222 patients. Most of the patients presented with respiratory (33.4%), GIT (14.9%), urological (14.7%) and skin (12.6%) complaints. The prescribers were either medical officers (MOs), clinical officers (COs) or nurses while the dispensers were either pharmacists or pharmaceutical technologists. Three facilities had neither a pharmacist nor a pharmaceutical technologist as the qualified dispensers at the facilities - dispensing was done by nurses. Cumulatively, 2636 drugs were prescribed to the out-patients in the 900 sampled prescription encounters. The majority of the prescribed drugs were analgesics/antipyretics (36.8%) and antibiotics (30.2%). The least prescribed drugs were antivirals (0.2%).

### Prescribing Indicators

The overall average number of drugs prescribed per patient encounter was 2.9 ± 0.5 (Prescribing indicator 1), ranging from one to eight drugs per prescription. The difference in the average number of drugs prescribed per patient encounter differed significantly between the 10 PPHCCs, *p* = 0.043. No facility had an average number of drugs prescribed that were within the WHO/INRUD recommended optimal range of 1.6 – 1.8.

Out of the 2636 prescribed drugs, 706 (27.7%) were written in their generic names (prescribing indicator 2); 1677 (63.6%) were prescribed by brand names and the remaining 253 (9.6%) had their generic names abbreviated. The practice of generic prescribing was observed to be significantly different among the PPHCCs, *p* = 0.005.

Out of the 900 prescription encounters, 795 (84.8%) had antibiotics (prescribing indicator 3). Amoxicillin was the most widely prescribed antibiotic followed by cotrimoxazole and metronidazole. The differences in antibiotic prescribing between the PPHCCs was statistically significant, *p* = 0.033.

Out of the 900 encounters, 224 (24.9%) included injections (prescribing indicator 4). The extent of prescribing of injections was statistically significant between the PPHCCs, *p* = 0.002. However, the percentage of prescriptions with the WHO/INRUD optimal value of 13.4% to 24.1% (15) for 7 of the 10 PPHCCs. Antipyretic and antibiotic injections were frequently prescribed. Diclofenac and Ceftriaxone injections were the most widely prescribed injection, at 38.2 and 24.4% respectively.

Out of the 2636 drugs prescribed, 2550 (96.7%) were prescribed from the KEML 2016 (prescribing indicator 5). The prescribing indicators are summarized in Table 1.

**Table 1:** WHO/INRUD Drug use indicators at Public primary healthcare centers in Kisii County, Kenya

| Indicators/PPHCC | Optimal value | 1 | 2 | 3 | 4 | 5 | 6 | 7 | 8 | 9 | 10 | Average | ANOVA |
| --- | --- | --- | --- | --- | --- | --- | --- | --- | --- | --- | --- | --- | --- |
| **Prescribing indicators** |  |  |  |  |  |  |  |  |  |  |  |  |  |
| Average drugs/ encounter | **1.6–1.8** | 2.6 ±.9 | 3.6 ± 1.0 | 3.0 ± 1.0 | 3.0 ± 1.0 | 3.6 ± 1.2 | 3.3 ± 1.1 | 3.0 ± .8 | 2.1 ± .0 | 2.6 ± .7 | 2.6 ± .8 | **2.9 ± .5** | 0.043 |
| % drugs by generic names | **100** | 12.7 | 17.4 | 46.1 | 19.4 | 14.8 | 21.4 | 7.8 | 13.7 | 69.1 | 54.3 | **27.7** | 0.005 |
| % antibiotic encounter | **20.0–26.8** | 84.4 | 91.1 | 77.8 | 83.3 | 82.2 | 84.4 | 90.0 | 86.7 | 84.4 | 83.3 | **84.8** | 0.033 |
| % injection encounter | **13.4- 24.1** | 3.3 | 50.0 | 16.7 | 10.0 | 67.8 | 35.6 | 24.4 | 10.0 | 21.1 | 10.0 | **24.9** | 0.002 |
| % drugs from KEML | **100** | 86.9 | 98.8 | 98.9 | 92.5 | 94.8 | 99.3 | 97.8 | 99.0 | 100.0 | 99.1 | **96.7** | 0.008 |
| **Patient-care indicators** |  |  |  |  |  |  |  |  |  |  |  |  |  |
| Average consultation time | **≥ 10 min** | 2.0 | 6.8 | 3.3 | 5.7 | 3.0 | 5.0 | 4.5 | 6.0 | 2.4 | 2.6 | **4.1** | 0.046 |
| Average dispensing time | **≥ 90 s** | 115.5 | 171.6 | 104.3 | 190.0 | 88.0 | 124.0 | 200.2 | 132.4 | 92.9 | 131.5 | **131.5** | 0.004 |
| % drugs dispensed | **100** | 86.7 | 55.0 | 73.1 | 78.5 | 61.8 | 84.5 | 79.3 | 86.2 | 72.0 | 85.9 | **76.3** | 0.001 |
| % drugs labeled | **100** | 10.8 | 29.5 | 36.7 | 93.5 | 25.0 | 8.5 | 1.4 | 17.9 | 4.5 | 3.0 | **22.6** | 0.002 |
| % patient knowledge | **100** | 44.7 | 60.0 | 55.5 | 46.7 | 53.3 | 60.0 | 71.1 | 56.7 | 50.0 | 46.7 | **54.7** | 0.005 |
| **Facility-specific indicators** |  |  |  |  |  |  |  |  |  |  |  |  |  |
| % copy of KEML | **100** | 100 | 0.0 | 100.0 | 0.0 | 0.0 | 0.0 | 0.0 | 0.0 | 0.0 | 0.0 | **20.0** | 0.005 |
| % key drugs available | **100** | 94.4 | 94.4 | 94.4 | 55.6 | 50.0 | 88.9 | 94.4 | 83.3 | 72.2 | 72.2 | **80.0** | 0.045 |

*1 = Oresi, 2 = Kegogi, 3 = Masimba, 4 = Entanda, 5 = Magena, 6 = Nyamagundo, 7 = Isecha, 8 = Egetuki, 9 = Kionyo, 10 = Mosoch*

### Patient-care indicators

The overall average consultation time for the 300 patients observed was 4.1 minutes (range 1 - 14 minutes) (patient – care indicator 1). The differences in consultation times among the PPHCCs was statistically significant, *p* = 0.046.

The average dispensing time was 131.5 seconds (range 45 - 360 seconds) (patient – care indicator 2). Again, the difference in dispensing times among the PPHCCs was statistically significant, *p* = 0.004.

Out of 872 drugs prescribed to the 300 recruited outpatients, 656 (76.3%) drugs were dispensed to the patients (patient – care indicator 3). Out of these 656 drugs dispensed to the outpatients, 148 (22.6%) were adequately labeled (Patient-care indicator 4). Majority of the dispensers only wrote the frequency of administration of drugs on the drug package or envelop/bag. WHO/INRUD recommends that each drug label should contain; patient name, dose regimen, dose, frequency of administration and quantity of the drug. (15).

The overall score on patients’ knowledge of drugs dispensed to them was 54.7% (Patient-care indicator 5). Patients’ knowledge of drug indications and dosage was good (77.0% and 75.7% of the patients correctly knew the indications and dosages of their drugs, respectively). However, very few patients (11.3%) were aware of the side effects of the drugs issued to them.

### Facility-specific indicators

Out of the 10 PPHCCs, only 2 (20%) reported having hard copies of the KEML 2016 booklets both at the prescribing and dispensing areas (Facility-specific indicator 1) There were no drug formularies available at any of the PPHCCs.

The availability of the 18 drugs selected from the KEML was assessed at the selected PPHCCs. Overall, 80.0% of the selected essential drugs assessed were available at the PPHCCs during the survey visit (Facility-specific indicator 2).

## DISCUSSION

Cumulatively, 2636 drugs were prescribed to the out-patients in all the 900 sampled prescription encounters. The majority of the prescribed drugs were analgesics/antipyretics 970 (36.8%) and antibiotics 795 (30.2%). The least prescribed drugs were antivirals (0.2%). The commonly prescribed analgesics were; paracetamol (43.7%), ibuprofen (19.4%), diclofenac (8.9%), and tramadol (5.2%).

The average number of drugs prescribed per prescription was 2.9. This was above the optimal range of 1.6 – 1.8 recommended by WHO/INRUD (15), indicating the likely practice of polypharmacy. In studies done in other countries, the average number of drugs per prescription was also higher than the recommended optimal range and ranged between 21.4 in Sudan (16), 2.5 in Egypt (14), 3.4 in Pakistan (17), 3.0 in Sri Lanka (18) and 4.8 in Ghana (19). Incompetent prescribers, unavailability of STGs, lack of continuous medical education (CME) programs and the unavailability of therapeutically potent drugs at the PPHCCs could be some of the reasons for the observed polypharmacy (17). Polypharmacy adversely influences patient treatment outcomes since patients are more likely to be noncompliant or experience adverse drug reactions (ADRs) (14). Rational prescribing is encouraged by the WHO/INRUD in order to avoid unnecessary excessive use (wastage) of drugs and probable adverse effects on the patients (17).

That percentage of drugs prescribed by their generic name was 27.7% indicating that clinicians attending to patients at the PPHCCs’ in Kisii County rarely prescribe drugs by their generic names. In studies carried out in other countries, the percentage of drugs prescribed by generic name was found to be exceedingly variable, from as low as 6% in Andorra (20) and 38.3% in Uzbekistan (21) to as high as 71.6% in Nigeria (8), 95.4% in Egypt (14) and 99.4% in Malawi (22). Previous studies done in Kenya had comparable findings, with 25.6% at Mbagathi District Hospital (23) and 45.5% at Makueni County Referral Hospital (24). The WHO/INRUD optimal percentage of drugs prescribed by the generic name is 100% (15). The findings of this study were way below the recommended value. This might be attributed to the belief of prescribers in branded drugs over generic products, extensive promotional activities by drug companies’ medical representatives to the prescribers or absence of a national policy of generic prescribing. The WHO/INRUD recommends prescribing drugs by their generic names. It gives clear identification, allows easy information exchange and allows improved communication among health professionals (17).

The percentage of encounters with antibiotics prescribed was 84.8%. The percentage was found to be higher compared to other studies. For instance, at Arba Minch and Chencha Hospitals in Ethiopia, the prevalence was 48.7% and 60.2% respectively (25). In India’s PHCCs, it was 60.9% (2),35.4% in Tanzania (26), 43.0% in Nepal (27), 33.1% in Burkina Faso (28), 50.0% in Burundi (29) and 28.8% in Brazil (29). The WHO/INRUD recommended value for percentage encounter with an antibiotic prescribed is 20 - 26.8% (15), suggesting that prescribers at the PPHCCs in Kisii County are overusing and misusing the antibiotics. The overuse and misuse of antibiotics leads to increased antibiotic resistance and wastage of scarce resources.

The percentage of encounters with an injection prescribed was 24.9%. This finding is comparable with the reported prevalence of injection prescribing of 27.6% at the PHCCs in Malawi (22) and 23.8% at Mbeya Health Center in Tanzania (26), and higher than the reported 3.0% in Nepal (27), 11.4% in Pakistan (17), 9% in Botswana (30) and 10.1% in Burundi (29). Other studies reported higher values such as 80.3% in Ghana (19) and 57.6% in Cambodia (29). The WHO/INRUD optimal value for percentage encounters with an injection prescribed is 13.4% to 24.1% (15). The prevalence of injection prescribing in this study (24.9%) was only slightly above the recommended range, which is encouraging. Antipyretic and antibiotic injections were frequently prescribed. Diclofenac and Ceftriaxone injections were the most widely prescribed injection, at 38.2 and 24.4% respectively.

The percentage of drugs prescribed from the KEML 2016 was 96.7%. All the PPHCCs had almost all the drugs prescribed from the KEML. This was higher than that reported in other previous studies conducted in Kenya - 72.2% at Mbagathi District Hospital (23) and 89.1% at Makueni County Referral Hospital (24). Other studies reported comparable findings, 95.4% in Egypt (14), 100.0% in Ethiopia (25), 96.7% in Tanzania (26) and 86.1% in Nepal (27). It was notable that though many PPHCCs in Kisii County did not have copies of KEML, they prescribed from the list. Prescribing drugs from the EML is one way of rational prescribing. However, prescribers may not choose drugs not in the EML due to the inadequate supply of EML copies (17).

The time that health – care providers devote to patients, majorly at the prescribing and dispensing service delivery points, determines the quality of disease diagnosis and management (25). The average consultation time was 4.1 min. The optimum WHO/INRUD value for average consultation time is ≥ 10 min (15). The time taken by the prescribers at the PPHCCs in the current study was shorter than that recommended to conduct a thorough patient assessment and prescribe drugs appropriately. This was comparable with findings reported in other countries where average consultation times ranged from 2.0 to 7.5 min (14) (17) (25) (27). However, the study conducted in Nigeria reported a better consultation time of 11.3 min (31). Insufficient consultation time can lead to an incomplete examination of patients and subsequently irrational therapy (32). Prescribers need to take sufficient time with patients in order to carry out comprehensive history taking, patient examination, provide suitable health education and ensure good clinician-patient rapport. This is significant as it ensures good patient-care. The increased workload of the prescriber and religious, ethnic or socioeconomic barriers between prescribers and patients could be the reasons for the short consultation time (17).

The average dispensing time was 131.5 seconds. The optimum value set by the WHO/INRUD for average dispensing time is ≥ 90 seconds (15). Based on the WHO/INRUD minimum time, the dispensers at the PPHCCs took sufficient time in processing the prescriptions and ultimately dispensing the prescribed drugs to the patients. In many studies conducted around the world, the average dispensing time was lower than that of the current study, ranging from 38 – 78 seconds (14) (17) (27) (31) (25). A study carried out at public hospitals in Ethiopia found more time taken by the dispensers at an average of 219.6 s (33). Adequately long dispensing time is required to explain key information about the drug(s) (dosage, adverse effects, and precautions) to the patient(s) as well as label the drug(s) adequately and dispense them to patients.

The percentage of drugs actually dispensed was 76.3%. The recommended optimal value of drugs actually dispensed by the WHO/INRUD is 100% (15). The finding of this study was less than those found in previous studies (14) (25) (31) (27). However, the percentage was higher compared to that reported at the public health facilities of Tanzania (56.2%) (26). The findings of this study could be an indication that some drugs may have been out of stock.

Drug labeling practice was very poor at the selected PPHCCs. The percentage of drugs dispensed adequately labeled was 22.6%. The poor labeling practices noted in this survey was similar to the findings of the survey performed at PHCCs in Eastern Province of Saudi Arabia (10.4%) (4) and Tanzania (20.1%) (26) where patient names and other vital details about the drug dosage regimen were not written in the labels (34). However, all drugs dispensed were adequately labeled (100.0%) in the Tertiary Care Hospital of India (35). The findings in Cambodia were worse (0.0%) compared to the current study (36). The omission of patient name, storage conditions and any other special precaution concerns on the drug label can lead to serious consequences such as drug misuse by patients (17).

Patients’ percentage knowledge on dispensed drugs was average, at 54.7%. The optimal WHO/INRUD value for patients’ percentage knowledge on correct drug dosage is 100% (15). The findings of this study (54.7%) was a little bit higher than from studies in India (46%) (35), Tanzania (37.9%) (26) and Malawi (27.1%) (22), but were much lower than those reported in Egypt (94.1%) (14), and Nigeria (93.2%) (31). Patient’s knowledge of drug dosage is important. It helps in improving patient care by avoiding the overuse of drugs and preventing ADRs/adverse effects that can cause harm to the patient’s health.

In any health - care center, availability of qualified prescribers and dispensers, adequate supply of key drugs and information access about drugs, such as EMLs/formulary, influences the ability to prescribe and dispense drugs rationally. Without these factors, it is difficult for healthcare workers to provide health services efficiently (15). Out of the 10 PPHCCs, only 2 (20.0%) had copies of the KEML 2016 booklets available both at the prescribing and dispensing areas. The findings were not consistent with the study carried out in Egypt where 8 (80.0%) out of 10 PHCCs had copies of the EML (14), 62.3% in Nigeria (31) and 67.4% in Malawi (22). The surveys in Nepal (27) and Pakistan (17) found that all the facilities (100.0%) had copies of EML. The WHO/INRUD requires that all health facilities have copies of EML (15). This is aimed at ensuring adherence of prescribers to the medicines listed in the EML when prescribing to promote the efficient provision of healthcare to patients (17).

Eighty percent (80.0%) of the selected essential drugs assessed were available at the PPHCCs at the time of the survey visit WHO/INRUD recommends 100% availability of essential drugs at the health facilities (15). The shortage of key drugs is detrimental to patients with regard to their health status and out-of-pocket expenses (15).

The use of WHO/INRUD guidelines on the three core drug use indicators and adherence to the WHO methodology offers more strength to the study. Also, adding to the study strength was; the use of a large sample size of 900 prescriptions and 300 outpatients.

The reasons for the irrational use of drugs could not be revealed in this study because it was limited. Further studies are necessary to disclose these reasons. Also, being a cross-sectional and retrospective study, there could have been an information bias and desirability.

## CONCLUSION

Most of the prescribing indicators greatly deviated from the WHO/INRUD recommended optimal values, indicating irrational drug use practices such as the practice of polypharmacy and misuse of antibiotics. Patient care and facility-specific indicators were also far from the optimal values except that of the average dispensing time. The findings of inadequately labeled drugs and poor patients’ knowledge of drugs dispensed to them were rather concerning

The County Health Management Team (CHMT) together with other stakeholders should implement interventions aimed at strengthening good prescribing and patient-care practices.

## Data Availability

The primary data gathered by the authors and which supports the findings of this study are available from the corresponding author upon request.

